# Longitudinal SARS-CoV-2 neutralization of Omicron BA.1, BA.5 and BQ.1.1 after four vaccinations and the impact of break-through infections in hemodialysis patients

**DOI:** 10.1101/2023.03.14.23287246

**Authors:** Louise Platen, Bo-Hung Liao, Myriam Tellenbach, Cho-Chin Cheng, Christopher Holzmann-Littig, Catharina Christa, Christopher Dächert, Verena Kappler, Romina Bester, Maia Lucia Werz, Emely Schönhals, Eva Platen, Peter Eggerer, Laëtitia Tréguer, Claudius Küchle, Christoph Schmaderer, Uwe Heemann, Oliver T. Keppler, Lutz Renders, Matthias Christoph Braunisch, Ulrike Protzer

**Affiliations:** Department of Nephrology, University Hospital rechts der Isar, Technical University of Munich, School of Medicine, Munich, Germany; Institute of Virology, Technical University of Munich, School of Medicine, Munich, Germany; TUM Medical Education Center, Technical University of Munich, School of Medicine, Munich, Germany; Max von Pettenkofer Institute & Gene center, Virology, Ludwig Maximilian University of Munich, Munich, Germany; Kidney Center Eifel Dialyse, Mechernich, Germany; KfH Kidney Center Harlaching, Munich-Harlaching, Germany; German Center for Infection Research (DZIF), Partner Site, Munich, Germany; KfH Kidney Center, Traunstein, Germany; Institute of Virology, Helmholtz Munich, Munich, Germany

**Keywords:** hemodialysis, SARS-CoV-2, COVID-19 vaccination, in-vitro viral neutralization, Omicron BA.1, BA.5, BQ.1.1

## Abstract

**Background:** Individuals on hemodialysis are more vulnerable to SARS-CoV-2 infection than the general population due to end-stage kidney disease-induced immunosuppression.

**Methods:** 26 hemodialysis patients experiencing SARS-CoV-2 infection after 3^rd^ vaccination were matched 1:1 to 26 out of 92 SARS-CoV-2 naïves by age, sex, dialysis vintage and immunosuppressive drugs receiving a 4^th^ vaccination with an mRNA-based vaccine. A competitive surrogate neutralization assay was used to monitor vaccination success. To determine infection neutralization titers, Vero-E6 cells were infected with SARS-CoV-2 variants of concern (VoC), Omicron sub-lineage BA.1, BA.5, and BQ.1.1. 50% inhibitory concentration (IC50, serum dilution factor 1:x) was determined before, four weeks after and 6 months after the 4^th^ vaccination.

**Results:** 52 hemodialysis patients received four COVID-19 vaccinations and were followed up for a median of 6.3 months. Patient characteristics did not differ between the matched cohorts. Patients without a SARS-CoV-2 infection had a significant reduction of real virus neutralization capacity for all Omicron sub-lineages after six months (p<0.001 each). Those patients with a virus infection did not experience a reduction of real virus neutralization capacity after six months. Compared to the other Omicron VoC the BQ.1.1 sub-lineage had the lowest virus neutralization capacity.

**Conclusions:** SARS-CoV-2-naïve hemodialysis patients had significantly decreased virus neutralization capacity six months after the 4^th^ vaccination whereas patients with a SARS-CoV-2 infection had no change in neutralization capacity. This was independent of age, sex, dialysis vintage and immunosuppression. Therefore, in infection-naïve hemodialysis patients a fifth COVID-19 vaccination might be reasonable 6 months after the 4^th^ vaccination.

## Introduction

Due to their impaired immune system hemodialysis patients are known to be at high risk for severe courses of COVID-19 ^1^. It has been observed that the currently available vaccinations against SARS-CoV-2 significantly reduce the mortality of COVID-19 in hemodialysis patients ^2^. Nevertheless, a lower humoral response to vaccination in hemodialysis patients compared to healthy controls has been reported as well as a significant decline in antibody levels and seropositivity over time ^3-5^. Meanwhile, the continuous emergence of new variants and subvariants with immune evasive properties ^6^ leads to an increased rate of break-through infections and necessitates research on the immune capacity generated by the initial wildtype specific vaccinations.

Hemodialysis patients are advised by German health authorities to perform a basic immunization consisting of three vaccinations followed by booster shots at least three months after the last immunization. Since homogeneous data on the impact of infections on immunity in hemodialysis patients is limited, there are however no clear recommendations for patients after SARS-CoV-2 infections ^7^.

An Omicron break-through infection in hemodialysis patients may induce higher Omicron specific antibody titers and less non-responders than vaccination only. A primary variant of concern (VoC) BA.1 or BA.2 infection though, seems to mainly generate variant specific humoral immunity ^8, 9^. In older healthy adults, break-through infections seem to generate a more durable humoral immunity than vaccinations alone ^10^.

The aim of this study was to investigate the course of the immune response in infection naïve compared to recently infected hemodialysis patients after the 4^th^ vaccination over 6 months independent of age, sex, dialysis vintage and immunosuppressive medication in an observational matched cohort study. Here, we present the results of the live-virus infection neutralization of SARS-CoV-2 VoC Omicron BA.1, BA.5, and BQ.1.1 and antibody-mediated immunity shortly before, after and at 6 months after the 4^th^ COVID-19 vaccination in a matched cohort of 52 hemodialysis patients.

## Material and Methods

### Study design

The COVIIMP study (German: “COVID-19-Impfansprechen immunsupprimierter Patient*innen”) is an observational cohort. The study design has been described before ^11^. In brief, the immune status and COVID-19 infections of hemodialysis patients and other immunocompromised patients are observed. The study conforms to the ethical guidelines of the Helsinki Declaration and has been approved by the local ethics committee (ethic vote 163/21 S-SR, March 19th, 2021, Medical Ethics Committee of the Klinikum rechts der Isar of the Technical University of Munich) and was reported (NIS592) to the Paul Ehrlich Institute. All study participants provided written informed consent.

### Study population

142 hemodialysis patients who had received four COVID-19 vaccinations until March 20^th^, 2022, were recruited from four dialysis centers (Kidney Center Eifeldialyse Mechernich, Germany, KfH Kidney Center München-Harlaching, Munich, Germany, KfH Kidney Center Traunstein, Traunstein, Germany and Klinikum rechts der Isar München, Munich, Germany). All patients were vaccinated by their treating physicians according to German guidelines. 20 patients were lost to follow up and 4 patients were excluded because they experienced a SARS-CoV-2 break-through infection already before the 3^rd^ vaccination resulting in a cohort of 118 patients. 26 of the 118 patients experienced a SARS-CoV-2 break-through infection after the 3rd vaccination and in the remaining 92 patients no history of SARS-CoV-2-infection was verifiable. Propensity score matching was carried out for the variables age, sex, dialysis vintage and presence of immunosuppressive medication using the MatchIt package in R version 4.2.2 (R Foundation for Statistical Computing, Vienna, Austria) to match these 26 patients with a history of SARS-CoV-2 infection to 26 out of 92 SARS-CoV-2-naïve patients (**Figure 1**).

**Figure 1.**
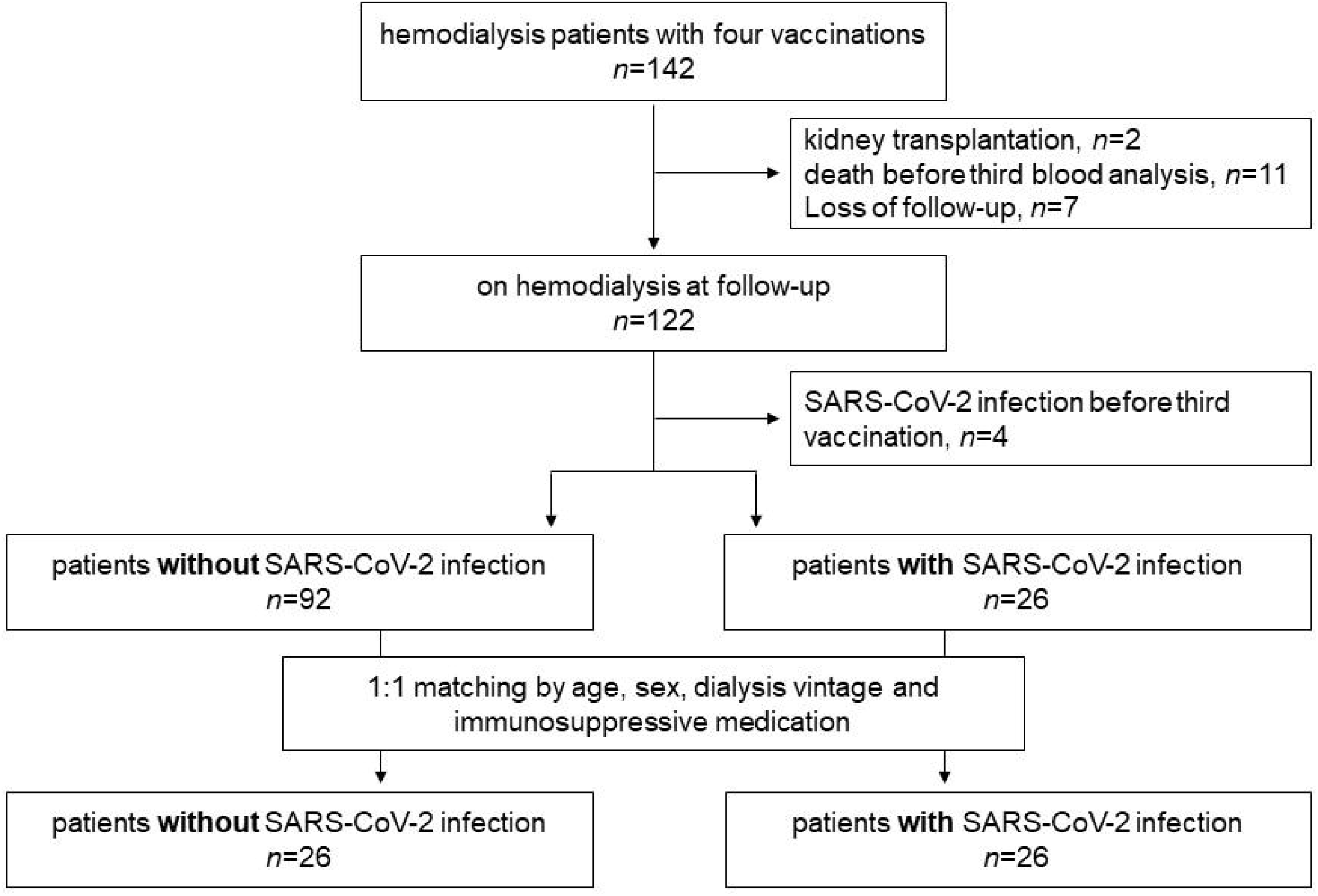
Flow chart of the study cohort. A cohort of 142 hemodialysis patients received a 4^th^ COVID-19 vaccination. 122 patients that could be followed up for six months (median 191 days) after 4^th^ vaccination on hemodialysis were enrolled in this study. At the end of follow-up, 30 out of the 122 patients (24.5%) had experienced a SARS-CoV-2 break-through infection. 26 patients were selected who experienced a SARS-CoV-2 infection after the 3^rd^ vaccination and were matched to SARS-CoV-2 naïve patients.

Blood analysis was performed three times in the 52 included participants between February and September 2022. The time of analysis was in median 124 (interquartile range [IQR] 103-124; minimum 27, maximum 133) days after the 3^rd^ vaccination, 26 (IQR 26-26; minimum 6, maximum 28) days, and 191 (IQR 191-192; minimum 161, maximum 201) days after the 4^th^ vaccination.

### SARS-CoV-2 infections

SARS CoV-2 infections were defined by a reported PCR-confirmed infection or by a new positive N-specific IgG level in the blood analysis. SARS-CoV-2 infections before the 2^nd^ blood sample were defined as early infection, break-through infections during the follow-up period were defined as late infections (**Figure 2**).

**Figure 2.**
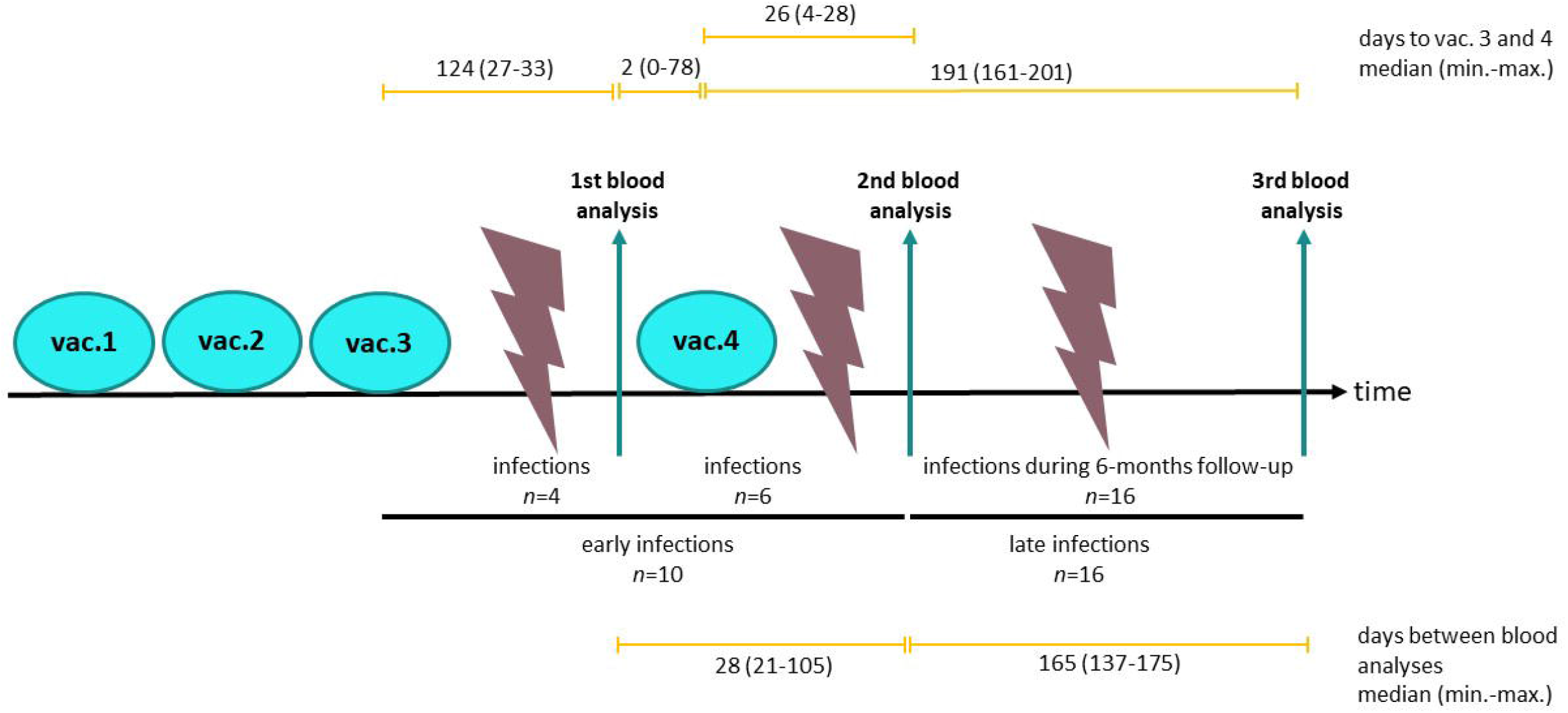
Timing of SARS-CoV-2 break-through infections. Patients with a SARS-CoV-2 infection after 3^rd^ vaccination but before the 2^nd^ blood sample taken in median 26 days after 4^th^ vaccination were defined as early infections (n=10). Patients with an infection during the follow-up period, i.e., after the 2^nd^ blood sample taken between in median 26 and 191 days after 4^th^ vaccination were defined as late infections (n=16). Abbreviations: vac. vaccination; min. minimum; max. maximum.

### SARS-CoV-2 IgG assay

Two types of SARS-CoV-2-specific IgG type antibodies were measured by chemiluminescent immunoassays (CLIA) using magnetic particle-based detection on an iFlash 1800 CLIA Analyzer (Yhlo Biotechnology, Shenzen, China) as described previously ^11^. The 2019-nCoV IgG kit (Yhlo) was employed for the detection of SARS-CoV-2 nucleocapsid specific antibodies (anti-N IgG) and considered positive at ≥ 10 AU/ml. SARS CoV-2 receptor-binding domain (RBD)-specific neutralizing antibodies (NAb) were measured using a surrogate neutralization assay (2019 nCOV NAb kit, Yhlo) that determines NAb titers preventing the binding of recombinant RBD (Wuhan strain) to the SARS-CoV-2 receptor ACE-2 protein. Titers were determined according to the WHO standard and are given in BAU/ml. Lower and upper limit of detection for NAb were 4 and 800 BAU/mL, respectively. For values exceeding the upper limit of quantification, a value of 801 AU/mL was puted into the statistical models.

### SARS-CoV-2 infection-neutralization assay

The method for analysis of patients’ serum neutralization capacity of several SARS-CoV-2 strains has been described in detail elsewhere^12^. Briefly, SARS-CoV-2 isolates of VoC Omicron BA.1 (B.1.1.529, GISAID EPI ISL: 7808190), VoC Omicron BA.5 (GISAID EPI-ISL: 15942298) and BQ.1.1 (GISAID EPI ISL: 15812430) were obtained from nasopharyngeal swabs of infected individuals. Vero E6 cells were incubated with the respective variants in DMEM (Dulbecco’s Modified Egale Medium) for two to three days and high titer virus stock was gained by collecting and centrifuging the supernatant. The viral stock was aliquoted and stored at -80 °C. Virus titers were determined by plaque assay before the start of analysis and viral strains were confirmed by next generation sequencing.

Patient’s sera were diluted 1:20 to 1:2560 and incubated for one hour with a defined multiplicity of infection of 0.03 plaque-forming units / cell (450 PFU/15,000 cells/well) at 37°C. Then, the inoculum was incubated for one hour on Vero E6 cells seeded into 96-well plates before removing the virus inoculum and washing the cells. After 24 hours, cells were fixed and permeabilized with 4% paraformaldehyde and 0.5% saponin buffer, blocked with 10% goat serum and stained with a primary anti-SARS-CoV-2-N antibody ((40143-T62, Sino Biological, Beijing, China). Afterwards, a colorimetric, quantitative analysis was performed by an in-cell ELISA using goat anti-rabbit IgG2a-HRP secondary antibody (12-348, EMD Millipore, Shanghai, China) and adding substrate TMB (tetramethybezidine).

After implementation of nonlinear regression, serum 50% inhibitory concentration (IC50) was defined as the dilution factor at which 50% infection inhibition was obtained. As described before ^11^, patients were classified as responders if the IC50 value of the infection neutralization was >1:20. Non-responders were defined for sera with a neutralizing IC50 value ≤1:20. Extremely strong responders exceeded the maximum detection range when diluted up to 1:40,960 (n=16). Graph Pad PRISM was used for calculations.

Analysis of neutralization capacity for VoC Omicron BA.1 before and after the 4^th^ vaccination had been performed and described previously ^11^ with the same protocol. Remaining samples were analyzed concurrently for BA.1 and analyses for BA.5 and BQ1.1 strains were added for all timepoints. Serum samples had been stored at -80°C after blood collection and were thawed and refrigerated at 4°C until analysis.

### Statistical Analysis

Variables are presented in frequencies and percentages, and as mean ± standard deviation (SD) (standard deviation), or median and interquartile range (IQR), as appropriate. The χ^2^ test or Fisher test was used for group differences, *t*-test and Mann Whitney U test were applied to continuous variables. Paired samples were tested with McNemar or Wilcoxon test, as appropriate. P-values <0.05 were considered significant, tests were carried out two-sided. Statistical Analysis was carried out with R version 4.2.2 (R Foundation for Statistical Computing, Vienna, Austria).

## Results

### Patient characteristics

Overall, 52 hemodialysis patients were included into the study (**Figure 1**). Patients were followed-up for a median of 6.3 (6.3-6.3) months after the 4^th^ vaccination. Patients had a median age of 71.4 (60.2-81.9) years. 9/52 patients (17.3%) were female. Median dialysis vintage at the time of the 4^th^ vaccination was 51.7 (24.2-94.9) months. One individual in each group received immunosuppressive medication, due to history of lung transplantation in the infection group (tacrolimus, mycophenolate mofetil, prednisolone) and due to anterior ischemic optic neuropathy (prednisolone) in the control group, respectively (**Table 1**). The matching variables age, sex, dialysis vintage and immunosuppressive medications did not differ between patients with a SARS-CoV-2 break-through infection compared to SARS-CoV-2-naïve patients (**Table 1**).

**Table 1.**
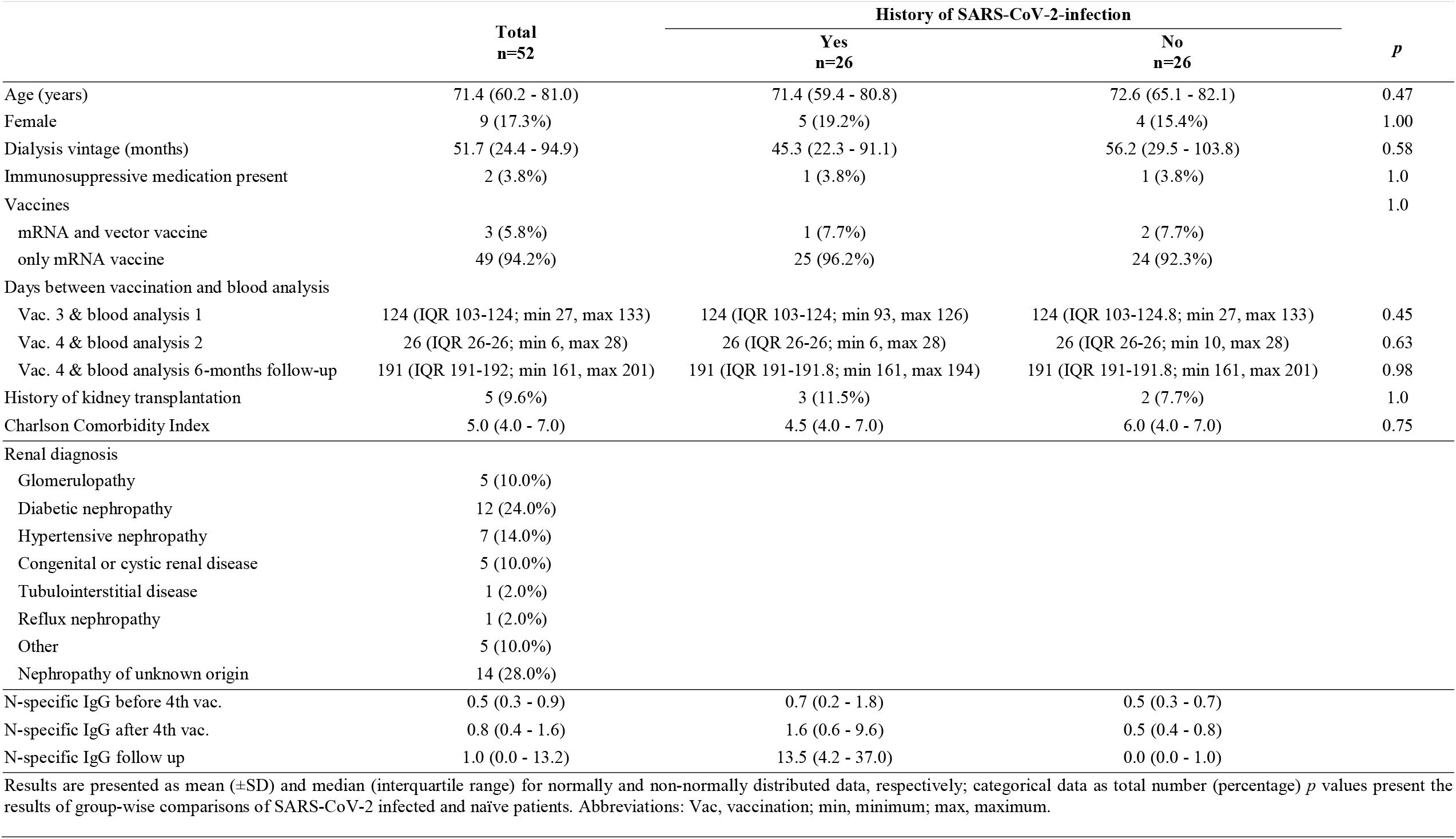
Patient characteristics.

### Vaccinations

Vaccinations were administered using mRNA vaccines (BNT162b2 by BioNTech-Pfizer or mRNA-1273 by Moderna) in 49 (94.2%) individuals. Three patients received a heterologous vaccination scheme (two patients received one shot AZD1222 by AstraZeneca and three shots of mRNA vaccines and one patient received two shots of AZD1222 by AstraZeneca and two shots of mRNA vaccines). Vaccination regimes did not differ between the two groups (p=1.0, **Table 1**).

### SARS-CoV-2 infections

SARS-CoV-2 break-through infections were detected in 26 patients after the 3^rd^ vaccination. Of these cases, 15 (57.7%) infections were PCR confirmed and 11 (42.3%) infections were identified by N-specific IgG-positivity only. Two patients were infected twice. In both cases the 1^st^ infection occurred before the 1^st^ vaccination.

In 10 of the 26 infected patients, the break-through infection occurred before the 2nd blood analysis, i.e., already before or till 26 days after 4^th^ vaccination. Of these, four (15.4%) patients were infected before the 1^st^ blood analysis (taken in median 4 months after 3^rd^ and 2 days before 4^th^ vaccination). Two of these infections were PCR confirmed and occurred 96 and 103 days before the 1^st^ blood examination. In the other six (23.1%) individuals the infection occurred after the 1^st^ and before the 2^nd^ blood examination taken in median 2 days before and 26 days after the 4^th^ vaccination, the PCR-confirmed infections (n=4) occurred in median 7.5 days after the 4^th^ vaccination.

Of the 26 infected patients, 16 (61.5%) had a SARS-CoV-2 break-through infection during the follow-up period i.e., 26 days to 6 months after 4^th^ vaccination (**Figure 2**). PCR confirmed infections (n=9) occurred in median 51 days before the 6-month follow-up blood analysis or in median 140 days after 4^th^ vaccination.

In line with this, N-specific IgG type antibodies were detectable in the infection group at the time of the 2^nd^ blood examination after the 4^th^ vaccination and in the follow up analysis (**Table 1**). Interestingly, no difference in serum neutralization titers after 3^rd^ and before 4^th^ vaccination (i.e at the 1^st^ blood examination) between naïve individuals and those experiencing a break-through infection were observed (**Table 2**).

**Table 2.** Group-wise comparison of immunity of hemodialysis patients with and without a recent SARS-CoV-2 infection.

|  | Total<br>n=52 | History of SARS-CoV2-infection |  | *p* |
| --- | --- | --- | --- | --- |
|  |  | Yes<br>n=26 | No<br>n=26 |  |
| **Neutralizing antibodies** |  |  |  |  |
| before 4th vac. (BAU/ml) | 717.5 (203.8 - ≥800) | 761.5 (165.5 - ≥800) | 455.0 (245.8 - ≥800) | 0.73 |
| after 4th vac. (BAU/ml) | ≥800 (≥800 - ≥800) | ≥800 (≥800 - ≥800) | ≥800 (≥800 - ≥800) | 0.92 |
| 6-months follow-up (BAU/ml) | ≥800 (417.2 - ≥800) | ≥800 (≥800 - ≥800) | 505.5 (232.8 - ≥800) | < **0.001** |
| **Omicron BA.1 infection neutralization capacity** |  |  |  |  |
| before 4th vac. (IC50) | 30.0 (0.0 - 195.8) | 48.5 (5.0 - 231.0) | 27.0 (0.0 - 141.2) | 0.49 |
| after 4th vac. (IC50) | 570.5 (198.0 - 2560.0) | 811.5 (167.0 - 2560.0) | 444.5 (222.2 - 2560.0) | 1.0 |
| 6-months follow-up (IC50) | 141.6 (20.0 - 663.6) | 619.2 (185.3 - 887.4) | 20.0 (0.0 - 20.0) | < **0.001** |
| **Omicron BA.5 infection neutralization capacity** |  |  |  |  |
| before 4th vac. (IC50) | 20.0 (20.0 - 116.1) | 22.5 (20.0 - 101.9) | 20.0 (20.0 - 113.7) | 1.0 |
| after 4th vac. (IC50) | 365.6 (76.1 - 2560.0) | 572.8 (107.6 - 2560.0) | 249.7 (62.7 - 2265.5) | 0.32 |
| 6-months follow-up (IC50) | 238.7 (20.0 - 2329.5) | 2339.0 (1041.5 - 7494.2) | 20.0 (20.0 - 74.9) | < **0.001** |
| **Omicron BQ.1.1 infection neutralization capacity** |  |  |  |  |
| before 4th vac. (IC50) | 20.0 (0.0 - 21.3) | 20.0 (0.0 - 20.0) | 20.0 (0.0 - 23.9) | 0.73 |
| after 4th vac. (IC50) | 99.0 (20.0 - 219.4) | 116.7 (20.0 - 255.7) | 89.1 (20.0 - 153.4) | 0.64 |
| 6-months follow-up (IC50) | 25.1 (20.0 - 266.3) | 270.0 (27.9 - 440.0) | 20.0 (0.0 - 20.0) | < **0.001** |
| Results are presented as median (interquartile range), *p* values present the results of group-wise comparisons of SARS-CoV-2 infected and naïve patients |  |  |  |  |
| Abbreviations: vac, vaccination |  |  |  |  |

Results are presented as median (interquartile range), *p* values present the results of group-wise comparisons of SARS-CoV-2 infected and naïve patients. Abbreviations: vac, vaccination

This indicates that the overall relatively low infection neutralization titers against the Omicron VoC BA.1, BA.5 and BQ.1.1 after three vaccinations are not sufficient to protect from infection with theses VoC

### Impact of a SARS-CoV-2 infection on immunity

All patients showed a significant increase in neutralizing antibody levels and neutralization capacity for the VoC Omicron BA.1, BA.5, and BQ.1.1 after a 4^th^ vaccination (**Figure 3, Table 2**). No significant differences, however, were seen between the two groups with or without a break-through infection neither at the 1^st^ blood analysis before nor at the 2^nd^ blood analysis in median 26 days after the 4^th^ vaccination (**Table 2**).

**Figure 3.**
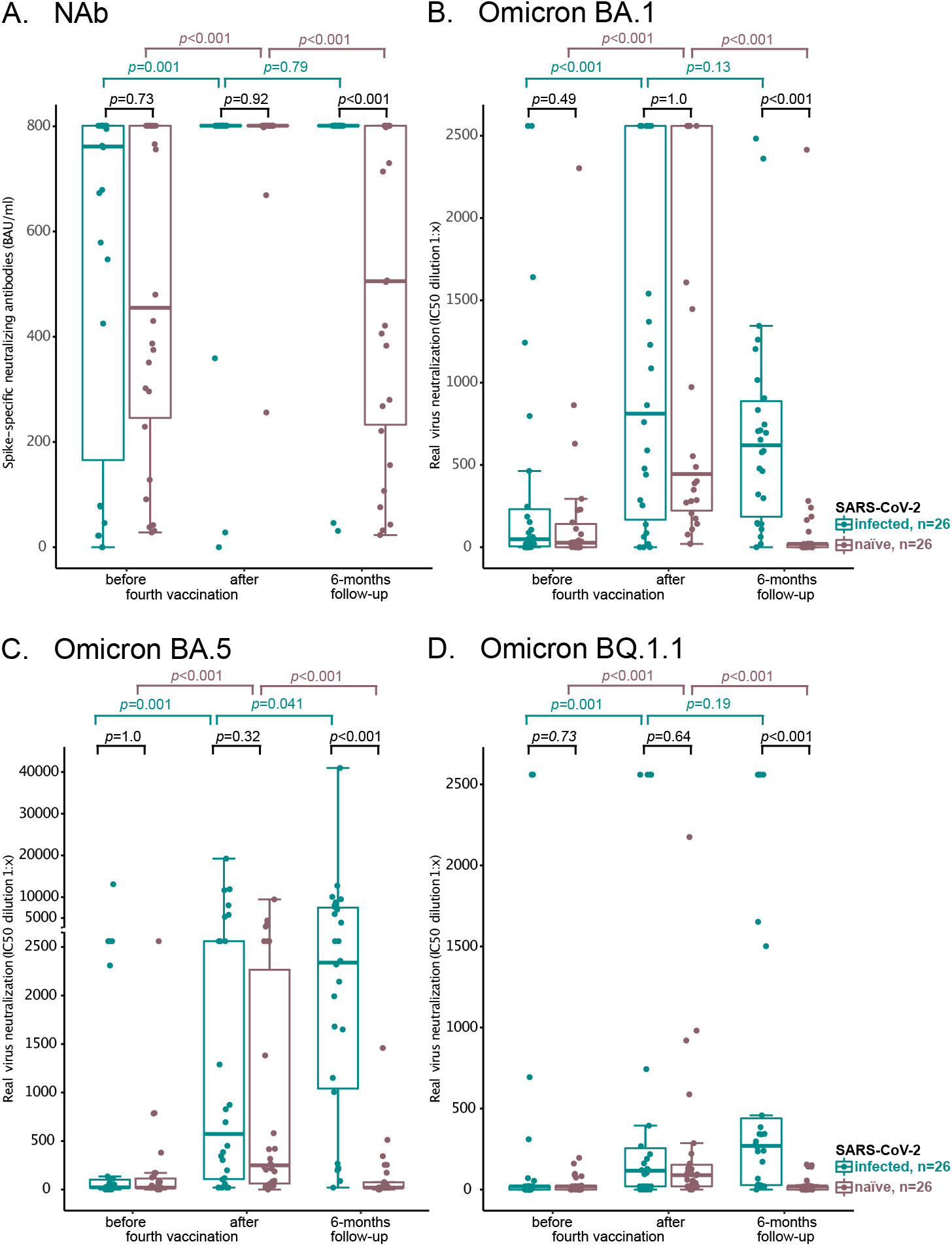
Neutralizing antibody titers before and after the 4^th^ vaccination and at 6-months follow-up. Hemodialysis patients who had experienced a break-through infection after 3^rd^ vaccination (infected) were compared to those without (naïve). In A) BAU/ml according to WHO standard using a competitive, surrogate neutralizing antibody assay are given. In B-D) 50% infection inhibition (IC50) titers in a real-virus neutralization assay with SARS-CoV-2 Omicron VoC BA.1 (B), BA.5 (C) und BQ.1.1 (D) are shown. Statistical analysis was performed using paired-samples Wilcoxon test, *p* values indicate statistical significance between groups. For better comparability the y-axis changes its linear scale at 2700 in C.

At the follow-up 6-months after 4^th^ vaccination, SARS-CoV-2 naïve patients had significantly lower virus neutralization capacities for all VoC (p<0.001) and significantly lower neutralizing antibody titers (p<0.001) (**Table 2, Figure 3**). Similar results were present when looking at the response rates (**Figure 4**). This indicates that a break-through infection confers a significant additional level of protection even against newly emerging VoC like BQ.1.1.

**Figure 4.**
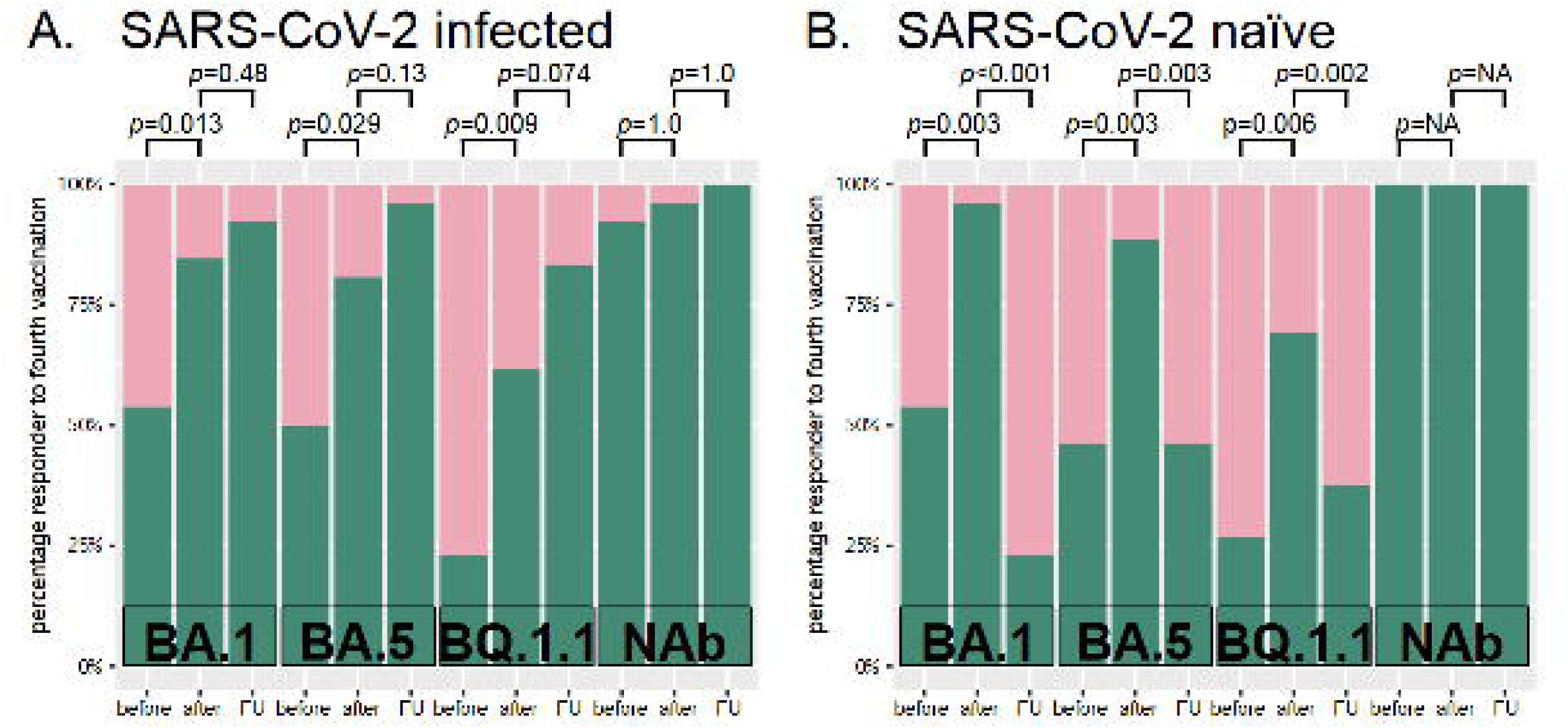
Percentage of vaccine responder before, after the 4^th^ vaccination and at 6-months follow-up. A) 26 hemodialysis patients who underwent SARS-CoV-2 break-through infection after 3rd vaccination were compared to B) 26 matched individuals which didn’t experience an infection. A responder was defined by Omicron BA.1, BA.5, and BQ.1.1 virus infection neutralization of ≥1:20 and as neutralizing antibodies (NAb) ≥10 BAU/mL. Green and red indicate the percentages classified as responder and non-responder, respectively. Statistical analysis was done using the McNemar test for paired samples. Abbreviations: NA, not applicable. FU, follow-up.

Comparing the virus neutralization capacity between the Omicron variants in the whole cohort we found significantly lower neutralization capacity of the Omicron BQ.1.1 variant compared to Omicron BA.1 or BA.5 directly after the 4^th^ vaccination (p<0.001) (**Figure 5**). At the six-months follow-up, neutralization capacity of Omicron BQ.1.1 and BA.1 was significantly lower compared to Omicron BA.5 (p=0.001 and p=0.013) (**Figure 5**). Stratification of virus naïve and infected patients showed similar results for both groups (data not shown).

**Figure 5.**
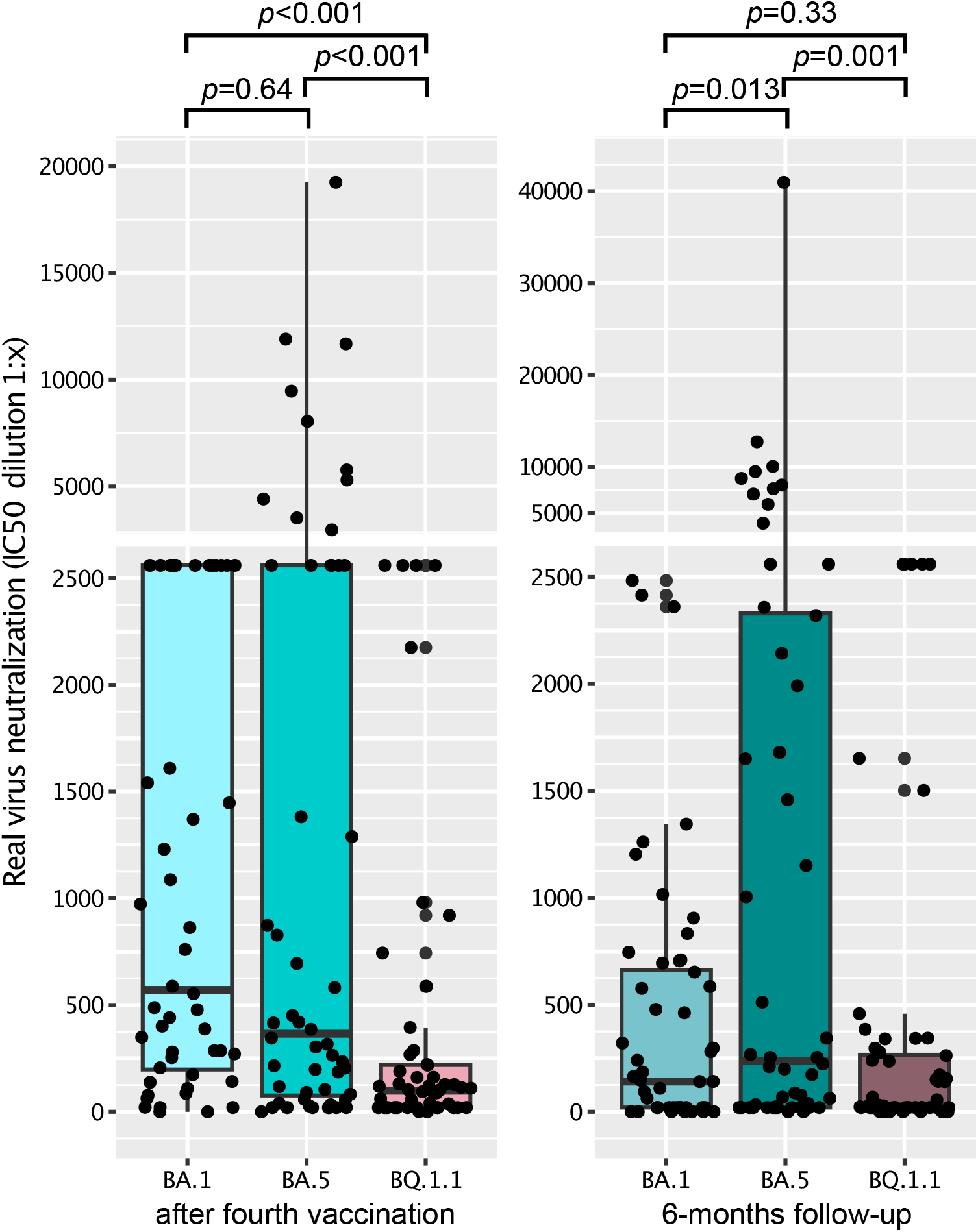
Comparison of neutralization capacity of Omicron variants BA.1, BA.5, and BQ1.1. in the whole cohort (infected and naïve patients (n=52)). Comparison of neutralization capacity after the 4^th^ vaccination (left side) and at the 6-months follow-up (right side). After the 4^th^ vaccination, neutralization capacity of the omicron variant BQ.1.1 was significantly lower than for BA.1 and BA.5. At the 6-months follow-up, neutralization capacity for BA.5 was significantly higher for BA.5 compared to BA.1 and BQ.1.1. Statistical analysis was performed using Wilcoxon test, *p* values indicate statistical significance between groups. For better clarity the y-axis changes its linear scale at 2700 in C.

Next, we compared hemodialysis patients with an early to those with a late SARS-CoV-2 infection **(Table 3, Figure 6). Figure 6** depicts serum neutralization titers against the different Omicron VoC and **Table 3** displays the median and interquartile ranges of the respective time points and provides a group comparison. Shortly after infection, i.e., at the 2^nd^ blood analysis time point for those with an early infection, we found significantly increased serum neutralization capacity for all VoC compared to those who had not yet (late infection) or not at all (naïve) experienced a SARS-CoV-2 break-through infection (**Table 3, Figure 6**). Patients with a later infection showed significantly improved neutralization capacity at the 6 months follow-up for Omicron variants BA.5 (p<0.001) and BQ.1.1 (p=0.002). Patients with an earlier infection had significantly decreasing serum neutralization capacity for Omicron BA.1 (p=0.004) and by trend slightly decreasing neutralization capacities for BA.5 (p=0.08) and BQ.1.1 (p=0.06) at the 6 months follow-up time point (**Figure 6**). However, despite this drop, neutralization titers remained significantly higher than in SARS-CoV-2-naïve hemodialysis patients (**Table 3, Figure 6**).

**Table 3.**
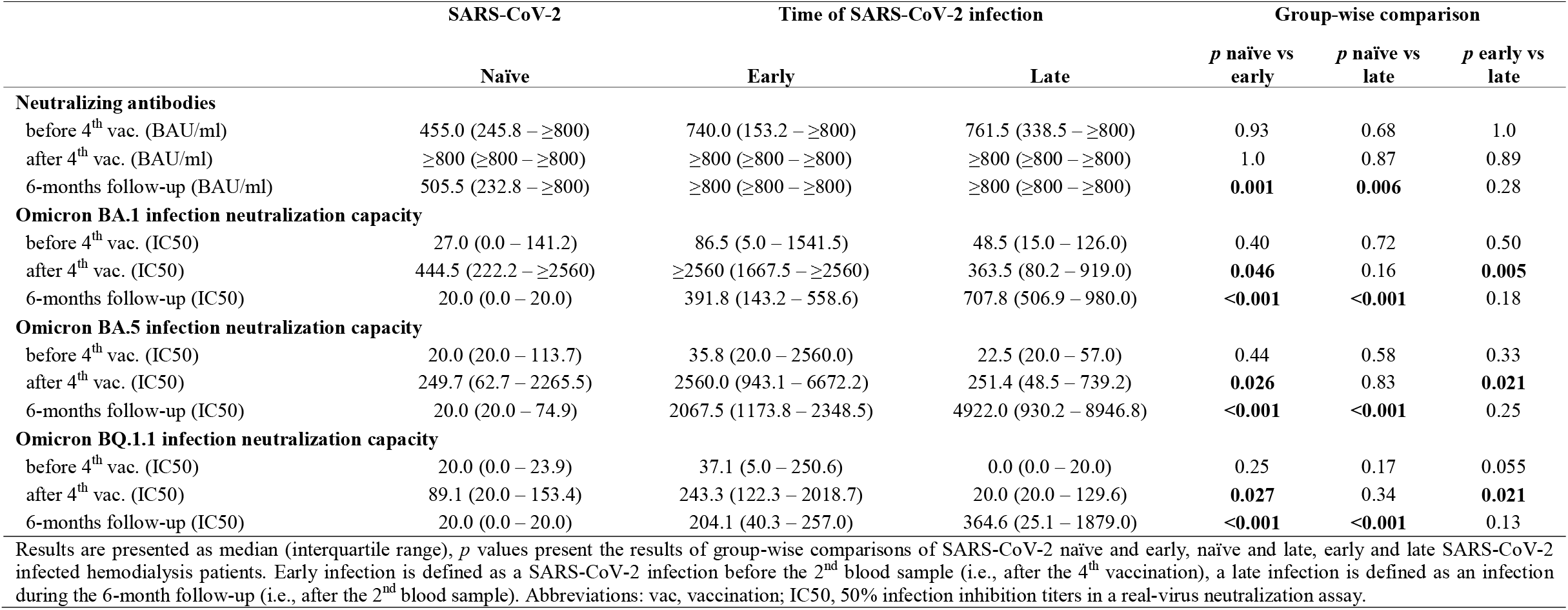
Group-wise comparison of neutralizing antibodies and neutralization capacity between SARS-CoV-2 infection naïve, early, and late infected individuals.

**Figure 6.**
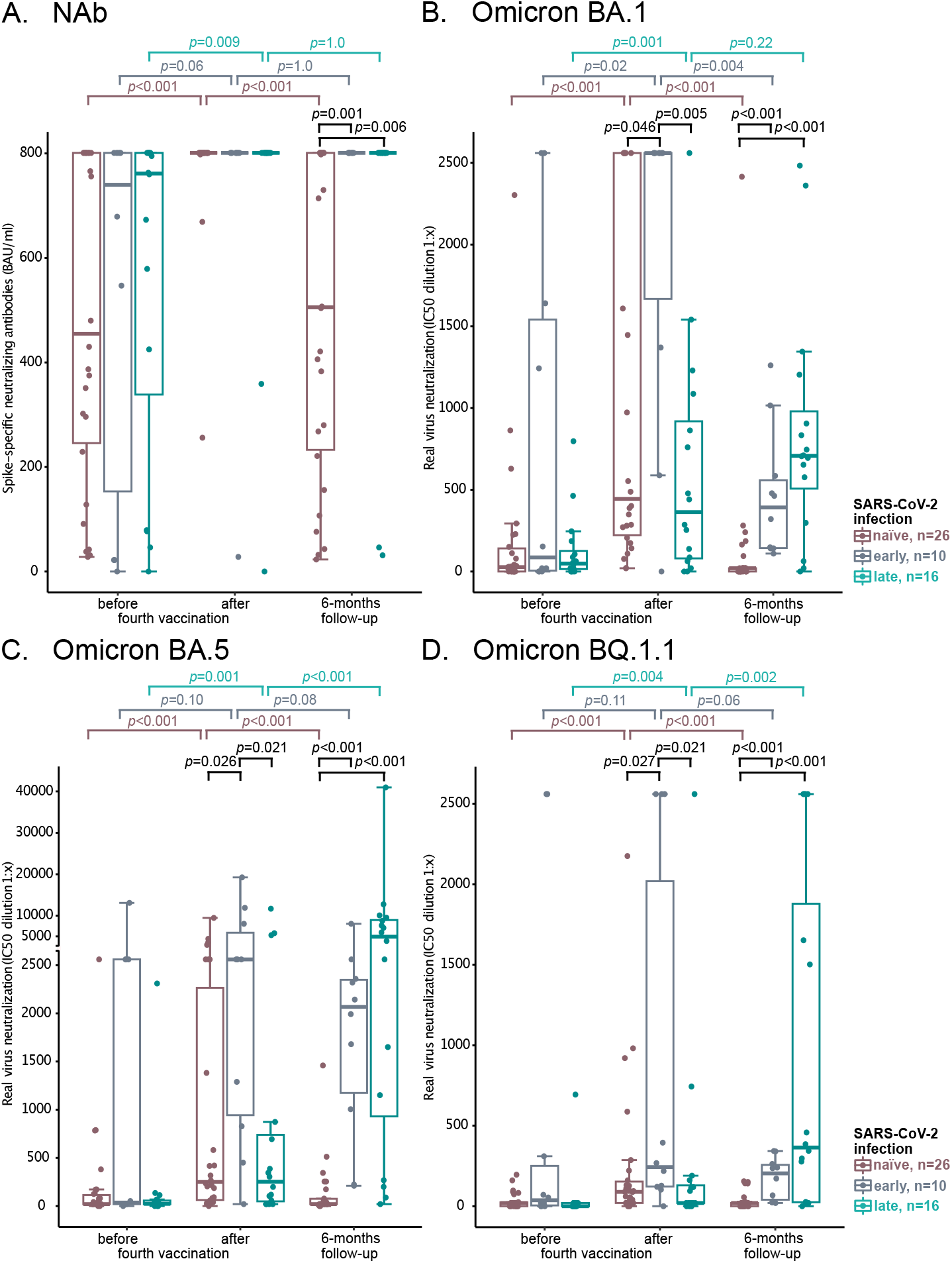
Neutralizing antibody titers before and after 4^th^ vaccination in infection-naïve patients and patients with SARS-CoV-2 break-through infection early and late after vaccination. Hemodialysis patients who had experienced a break-through infection after 3^rd^ vaccination until median 26 days after 4^th^ vaccination were grouped as early infection (early) and compared to those experiencing no infection and those experiencing an infection after the 2^nd^ blood examination taken between in median 26 and 191 days after 4^th^ vaccination (late). In A) BAU/ml according to WHO standard using a competitive, surrogate neutralizing antibody assay are given. In B-D) 50% infection inhibition (IC50) titers in a real-virus neutralization assay with SARS-CoV-2 Omicron VoC BA.1 (B), BA.5 (C) und BQ.1.1 (D) are shown. Statistical analysis was performed using unpaired and paired-samples Wilcoxon test, *p* values indicate statistical significance between groups. For unpaired analyses only significant *p* values are shown. For better comparability the y-axis changes its linear scale at 2700 in C.

## Discussion

This prospective matched observational study demonstrates that virus neutralization capacity for all current VoC has decreased significantly in SARS-CoV-2 naïve hemodialysis patients compared to SARS-CoV-2 infected patients six months after the 4^th^ vaccination independent of age, sex, dialysis vintage and immunosuppression. The strength of our study is the examination of the live-virus infection neutralization capacity of patients’ sera for different SARS-CoV-2 Omicron VoC. Compared to the other Omicron VoC BA.1 and BA.5, the neutralization capacity of the late Omicron BQ.1.1 variant was the lowest. This was true directly after the 4^th^ vaccination as well as at the 6-months follow-up. These findings are highly important, because Omicron BA.5 has been the predominant VoC in Germany during the follow-up period of this study and Omicron BQ.1.1 is currently (as of February 2023) the most frequent VoC in Germany with increasing prevalence^13^. It will therefore be important to further closely monitor new variants that might escape the immune system or erase the effectiveness of established antibody treatments ^6^.

We previously showed that by administration of a 4^th^ vaccine dose a significant increase of the virus neutralization capacity can be achieved for VoC Delta and Omicron BA.1 ^11^. With this work we add that this also holds true for VoC Omicron BA.5 and BQ.1.1. As we found significantly decreasing virus neutralization capacity in the SARS-CoV-2 naïve group for all observed Omicron variants during the follow-up period of six months, we assume that the humoral immunity provided by the vaccines that were designed for the original SARS-CoV-2 wildtype strain is rapidly waning over time and in their ability to neutralize current Omicron VoC. In SARS-CoV-2 naïve hemodialysis patients a fifth vaccination 6 months after the last vaccination could be a reasonable approach – ideally using an adapted vaccine. During the observation period of our study a specific Omicron adjusted vaccine was not available. Today, an Omicron specific vaccine has become available that could further improve the immune response^14^.

In SARS-CoV-2 infected patients no decreased neutralization capacity was observed at six months follow up for VoC BA.1 and BQ.1.1, and we found a significantly higher neutralization capacity for Omicron BA.5. This might be explained by the fact that most patients in our cohort were infected when the Omicron BA.5 VoC was by far the dominant variant. However, it remains open if the number of exposures to the viral spike protein or the type of spike involved in the booster was the relevant difference. In our study, the infected patients had five exposures while SARS-CoV-2 naïve patients had only four and were “only” exposed to first-generation vaccines. Alternatively, exposure to a different type of spike protein during break-through infection with the current Omicron variants could be the key for a broader immunity against the latest VoC. However, the effect of time has to be considered, since the follow-up analysis in patients with a late infection was closer to the virus exposure than in patients with an early infection or no history of infection. When comparing patients with an early infection to those without a history of infection we still found an improved neutralization capacity at the follow-up in infected patients, indicating that patients might benefit from a fifth exposure to an Omicron spike protein independently of time (**Table 3, Figure 6**). The increased breadth of immunity observed would argue for using an Omicron BA.5 adapted vaccine.

Overall, we were surprised that only 25% of our hemodialysis cohort had a SARS-CoV-2 infection until the end of our observation period. Additionally, the infection was asymptomatic in 42% of the cases and diagnosed by N-specific IgG-positivity only. This high rate of asymptomatic infections could be explained by the fact that all patients had received at least three COVID-19 vaccinations at the time of the SARS-CoV-2 infection. We did not record the variants, however as most infections occurred after mid-March, Omicron infections are most likely since initially Omicron BA.1 and BA.2 and from June onwards Omicron BA.5 were the predominant variants in Germany ^13, 15, 16^. Interestingly, in patients with late infections (after the 2^nd^ blood examination) a significantly increasing neutralization capacity could only be found for VoC BA.5 and BQ.1.1 during the follow-up period. This could be due to a relevant proportion of patients which might have been infected with the VoC BA.5 during the follow-up period. A break-through infection with BA.5 might therefore also positively impact and broaden immunity against the currently predominant variant BQ.1.1 which is a variant that was not present in Germany during the study period while it doesn’t seem to impact immunity against Omicron BA.1. Furthermore, it argues for using an Omicron BA.5-adapted vaccine for booster vaccinations.

Low serum anti-spike concentrations have been associated with an increased risk of break-through infections in hemodialysis patients ^17, 18^. We detected high surrogate NAb titers against the original Wuhan-strain in all our patients although absolute NAb values were significantly decreasing in SARS-CoV-2 naïve patients during the follow-up. It is unknown which NAb level might be an appropriate surrogate cut-off level to evaluate if a patient needs a booster COVID-19 vaccination, especially, when these surrogate neutralizing antibody measurements are designed to detect wildtype specific antibodies.

Finally, some limitations must be mentioned. We have focused our analyses on the most relevant VoC for Germany. Our results are not generalizable to other variants such as the recently described Omicron XBB.1.5 VoC with emerging prevalence in the United States. This variant has been associated to immune escape and ineffectiveness of established antibody treatments ^6^ and even seem to have additional immune escape potential when compared to the BQ1.1 VoC^19^. However, a more detailed analysis in patient cohorts is still lacking.

## Conclusion

In conclusion, we found a significantly reduced immune response against the recent SARS-CoV-2 VoC in hemodialysis patients. Low serum neutralization capacity within three months after a 3^rd^ vaccination could not prevent Omicron break-through infection. In addition, we found a rapid waning of immunity within 6 months after a 4^th^ vaccination in SARS-CoV-2 naïve patients which was not detected after an Omicron break-through infection. Therefore, another booster vaccination with an Omicron-adapted vaccine seems reasonable in hemodialysis patients without a SARS-CoV-2 infection. Furthermore, close monitoring of circulating SARS-CoV-2 variants will be necessary to identify those conferring an increased risk for hemodialysis patients.

## Key learning points

### What is already known about this subject?

- Hemodialysis patients are more likely to experience a severe course of COVID-19.
- Immunity after vaccination is less pronounced and diminishes more quickly in hemodialysis patients compared to healthy controls.
- Break-through infections broaden immunity in the general population.

### What this study adds?

- A 4^th^ vaccination with the original SARS-CoV-2 wildtype-based vaccine induces humoral immunity against currently predominant Omicron variants of concern BA.5 and BQ.1.1. However, compared to the other Omicron VoC patients’ sera showed the lowest virus neutralization capacity of the currently dominant Omicron VoC BQ.1.1 sub-lineage and immunity against all VoC wanes 6 months after 4^th^ vaccination.
- The overall low neutralizing antibody titers in hemodialysis patients 4 months after 3^rd^ vaccination were not sufficient to prevent infection with Omicron VoC with high immune escape capacity.
- Exposure to a different SARS-CoV-2 spike protein during break-through infection broadens immunity in hemodialysis patients and renders it more sustained.

### What impact this may have on practice

- The use of a vaccine adapted to newer SARS-CoV-2 variants should be of advantage for hemodialysis patients.
- Fourfold vaccinated SARS-CoV-2 naïve patients should consider a fifth vaccination six months after the last vaccination.
- Patients experiencing an Omicron break-through infection after vaccination possess a broader humoral immunity against current variants than patients vaccinated only.

## Data Availability

The datasets for this manuscript are not publicly available because written informed consent did not include wording on data sharing (German data protection laws). Reasonable requests to access the datasets should be directed to the corresponding author.

## Authors’ Contributions

LP, BH, LR, UP, and MCB wrote the 1^st^ draft of the manuscript. LP and MCB performed the statistical analysis. LP, MT, VK, CHL, MW, ES, EP, PE, LT, CK, CS contributed to blood sampling and data acquisition. CCC, CC, CD, RB, OTK, BHL performed in vitro virus neutralization assays and measurement of neutralizing antibodies. Project supervision was done by LR, UH, UP, and MCB. Each author contributed important intellectual content during manuscript drafting or revision and accepts accountability for the overall work by ensuring that questions pertaining to the accuracy or integrity of any portion of the work are appropriately investigated and resolved.

## Institutional Review Board Statement

The study, conforming to the ethical guidelines of the Helsinki Declaration, was approved by the Medical Ethics Committee of the Klinikum rechts der Isar of the Technical University of Munich (approval number 163/21 S-SR, 19 March 2021) and registered at the Paul Ehrlich Institute (NIS592).

## Acknowledgements

We would like to thank all patients for their participation in the study.

